# Reliability and Quality Assessment of Internet Videos as Guidance for Dietary Weight Loss Intervention: A Cross-Sectional Study in China

**DOI:** 10.1101/2025.05.15.25327656

**Authors:** Xiaohong Chen, Shiqi Zhou, Yongjian Zhang, Qianqian Zhong, Fengqin Sun, Rui Xu, Zhongli Sun, Junying Sun, Lin Yang, Shiqi Yan, Qianle Zhang, Zhuoxian Xie, Yongjiang Cai, Yanwu Xu, Hanyi Yu

## Abstract

**Background:** Obesity, a chronic disease affecting nearly all physiological functions, poses severe public health challenges by increasing the risk of conditions like diabetes, cardiovascular diseases, and depression. Managing obesity typically involves altering energy balance, with dietary interventions emerging as an effective and sustainable weight management approach. As internet usage surpasses 4 billion globally, many individuals now turn to online platforms for health information, with video content on platforms like TikTok gaining significant popularity for its accessibility and engagement. However, the quality of such content varies widely. In China, platforms like TikTok, BiliBili, and Kwai dominate health information dissemination, yet limited studies have evaluated the reliability of their dietary weight loss video content.

**Objective:** Our study aims to assess the reliability and quality of the information in Chinese videos on dietary weight loss shared on the BiliBili, TikTok, and Kwai, three video–sharing platforms.

**Methods:** We collected the top 100 dietary weight loss videos on BiliBili, TikTok, and Kwai in February 2024 and evaluated the information quality and reliability of the videos using the Global Quality Score (GQS) and mDISCERN. We also analyzed the correlation between video quality and video characteristics.

**Results:** The average GQS scores for dietary weight loss videos on BiliBili, TikTok, and Kwai are 2.04, 1.81, and 1.7, respectively, while the average mDISCERN scores are 2.01, 1.81, and 1.73. The median scores for both GQS and mDISCERN across the three platforms are 2. Although BiliBili’s GQS and mDISCERN scores are higher than those of Kwai and TikTok (GQS: P<0.01, P=0.02; mDISCERN: P=0.02, P=0.08), none of the platforms have exceeded a score of 3, indicating that the quality and reliability of dietary weight loss videos on BiliBili, TikTok, and Kwai are all quite poor.

Additionally, our study identified several significant positive correlations: the GQS score was positively correlated with video duration (r=0.41, P<0.01), as was the mDISCERN score (r=0.32, P<0.01). Strong positive correlations were also observed between the number of likes and favorites (r=0.90, P<0.01), likes and comments (r=0.92, P<0.01), as well as favorites and comments (r=0.86, P<0.01).

**Conclusions:** Our research found that although the GQS and mDISCERN scores of BiliBili were higher than those of TikTok and Kwai, the final scores of the three platforms did not exceed 3 points, indicating that the video quality of the three diet video websites was low. Video social media platforms should establish relevant policies to supervise and review the publication of medical science popularization videos, providing reliable sources of information for public health.

## Introduction

Obesity is a chronic disease, like hypertension and asthma, which has almost adverse effects on all physiological functions of the human body. The World Health Organization defines overweight and obesity as abnormal or excessive fat accumulation that poses a threat to health. BMI is one of the indicators used to measure the degree of body obesity, which is calculated by dividing a person’s weight (in kilograms) by the square of their height (in meters) [1]. About 2000 years ago, people began to pay attention to the impact of obesity on incidence and mortality rates [2]. A substantial body of research has demonstrated that obesity increases the risk of developing diabetes [3], cardiovascular diseases [3, 4], or psychological disorders such as depression [5]. Obesity is a serious public health threat. The core principle of any obesity treatment is to alter the balance between energy intake and energy expenditure, ensuring that energy expenditure exceeds energy intake. Medicinal interventions and bariatric surgery are common weight loss methods [6], but they require considerable manpower and resources and are often inefficient and costly. However, a growing body of evidence suggests that dietary intervention is a critical factor in short-term and long-term weight loss [7–9], and can achieve ideal weight management without compromising health. Therefore, it is crucial for the public to assess the reliability of dietary intervention information gathered in daily life for weight loss.

With the advancement of science and technology, the way people access medical information has undergone significant changes. Compared to traditional methods such as consulting doctors or reading medical books, more and more individuals prefer to seek medical information online [10,11]. As of 2021, the number of global Internet users has exceeded 4 billion [12], and studies have found that one-third of American adults use the Internet to learn about health issues [13]. In recent years, video information has become increasingly popular. Statistics show that videos with the hashtag #weightloss on TikTok have been viewed more than 9.7 billion times worldwide [14], indicating that healthy weight loss is a global hot topic. Compared to traditional text information, social media videos have the following advantages: firstly, visual information is more easily absorbed and remembered than text, and can quickly and inexpensively provide health information to a wide audience; secondly, videos encourage users to engage in healthy behaviors spontaneously through their rich visual effects [15,16]. However, due to the large number of content creators on video social platforms and less regulation of video content, the quality of information on diet and weight loss online is uneven, and people may inadvertently encounter incomplete or misleading information [17,18]. Therefore, quality control of medical science popularization videos online is essential.

In China, BiliBili, TikTok (Chinese version also named Douyin), and Kwai (Chinese version also named Kuai Shou) are three most popular video-sharing platforms.

These platforms are currently the main online media for disseminating health information online, with a monthly activity level exceeding 100 million people [19]. All information on these platforms is freely accessible and users only need to search for keywords of interest to view related videos. Previous studies have evaluated the quality of popular medical science videos on different topics on TikTok and BiliBili. Videos related to plastic surgery are considered reliable [20], while the quality of videos on liver cancer and gallstones is generally unsatisfactory [21, 22]. Most current research focuses on analyzing the quality of videos related to weight loss on English-language platforms such as YouTube and TikTok [23,24]. However, to the best of our knowledge, there has been no research analyzing the quality and reliability of videos dietary weight loss intervention on video-sharing platforms in China. In response to the gap in the research literature, the purpose of this study is to assess the quality of the most popular dietary weight loss videos on BiliBili, TikTok, and Kwai.

## Methods

### Search Strategy and Data Collection

As shown in Figure 1, in this cross-sectional study, we searched for videos related to “dietary weight loss” on BiliBili (Chinese version 7.68.0), TikTok (Chinese version 28.6.0), and Kwai (Chinese version 12.0.40) on February 28th, 2024. To mitigate biases in video recommendations stemming from the personalized algorithms of the platforms, we conducted searches using newly created accounts on each platform without imposing any additional constraints. We then ranked the videos based on the algorithms of the respective platforms and, after eliminating videos unrelated to “ dietary weight loss” and those with identical content from various sources, we selected the top 100 videos as our primary data set.

**Figure 1.**
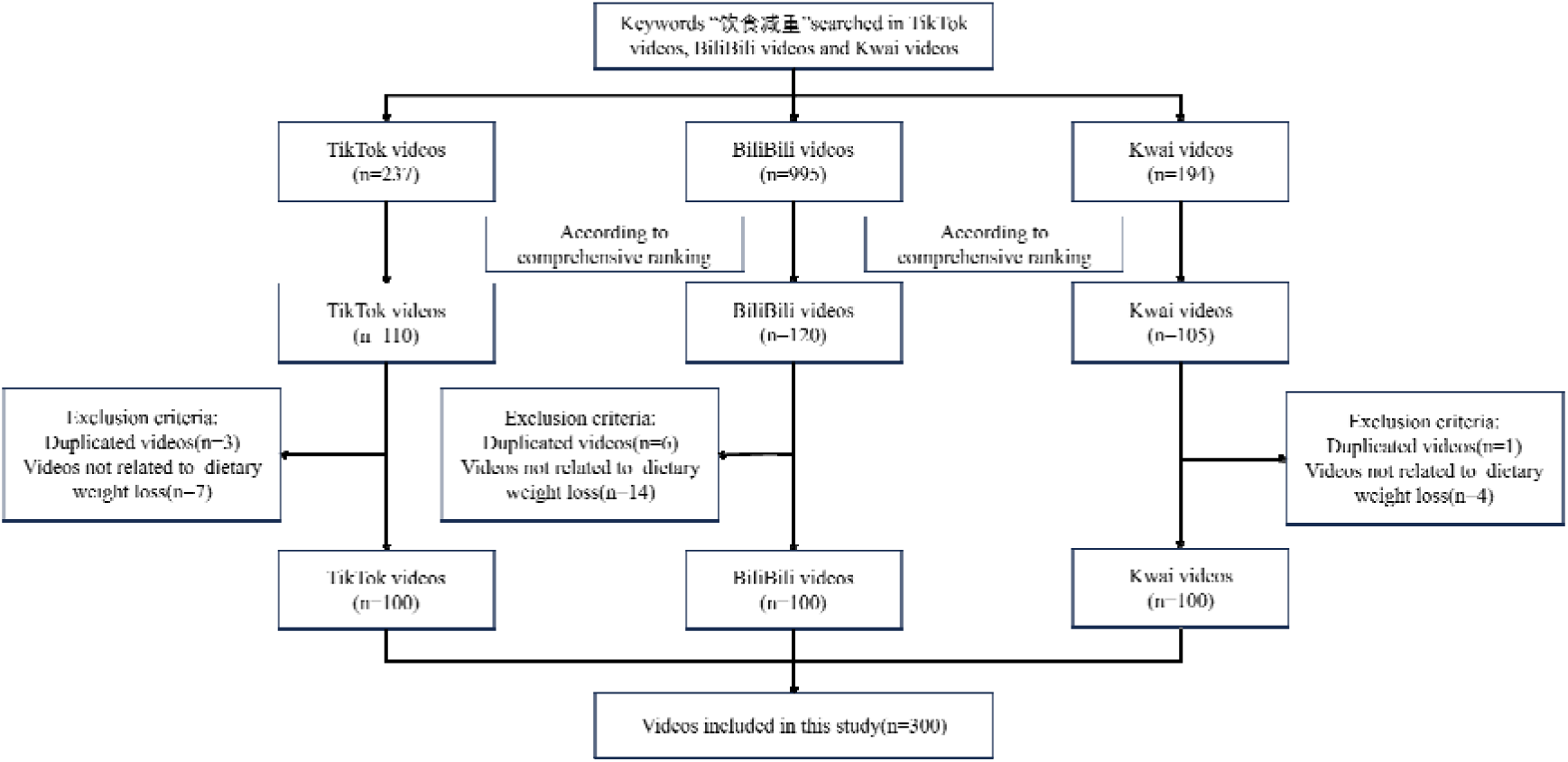
Search strategy for videos on dietary weight loss.

To ensure the credibility of the video content, we meticulously verified the details of the video publishers. For instance, if a publisher was purported to be a medical professional from a hospital, we confirmed the accuracy of this information on the hospital’s official website. Beyond this, we also gathered ancillary reference information for the videos, including the video’s title, the publisher’s name, duration, and the count of current likes and comments. All collected data were securely stored in Excel (Microsoft Inc.) for further analysis.

We categorized the videos into two groups based on their publishers: professional doctors and non-doctors. Additionally, the videos were classified into four content types: personal experiences, weight loss knowledge, advertisements, and other categories.

### Ethical Considerations

This study did not utilize any clinical data, human samples, or laboratory animals. All data were sourced from publicly available videos on BiliBili, TikTok, and Kwai, ensuring no personal privacy issues were involved. As the study did not engage directly with users, it did not require ethical review.

### Video Assessment

#### Assessment Criteria

We utilized the mDISCERN and the Global Quality Score (GQS) tools to assess the reliability and informational quality of the videos, respectively, the validity of which has been confirmed in previous studies [25–27]. The mDISCERN is an adaptation of the DISCERN instrument proposed by Singh et al., and it evaluates the reliability of videos across five dimensions: clarity, relevance, fairness, stability, and plasticity [25,28]. Each dimension is scored individually, with a total score ranging from 0 to 5; the higher the score, the better the reliability of the video. Compared to DISCERN, mDISCERN demonstrates improved convenience and accuracy in assessing video quality [29]. The GQS, introduced by Bernard et al., was initially designed to evaluate the quality of health information on websites and has since been widely adopted for assessing the quality of video information [26,30]. The GQS systematically evaluates the quality of videos based on informational quality, traffic, and usefulness, with a scoring range from 1 (very poor) to 5 (excellent) [30].

Detailed scoring criteria for the GQS and mDISCERN are presented in Table 1 and Table 2.

**Table 1.** mDISCERN scoring standard.

| mDISCERN | Description |
| --- | --- |
| Question 1 | Is the video clear, concise, and understandable? |
| Question 2 | Are valid sources cited? |
| Question 3 | Is the content presented balanced and unbiased? |
| Question 4 | Are additional sources of content listed for patient reference? |
| Question 5 | Are areas of uncertainty mentioned? |

**Table 2.** GQS scoring standard.

| Level | Description |
| --- | --- |
| 1 | Poor quality, poor flow of the site, most information missing, not at all useful for patients |
| 2 | Generally poor quality and poor flow, some information listed but many important topics missing, of very limited use to patients |
| 3 | Moderate quality, suboptimal flow, some important information is adequately discussed but others poorly discussed, somewhat useful for patients |
| 4 | Good quality and generally good flow, most of the relevant information is listed, but some topics not covered, useful for patients |
| 5 | Excellent quality and excellent flow, very useful for patients |

#### Assessment Methods

We compiled the collected original videos and pertinent information, and then dispatched them to the professional raters involved in the scoring process. All participating video raters had extensive clinical expertise in dietary weight reduction. To minimize scoring discrepancies, every rater thoroughly reviewed the detailed criteria for GQS and mDISCERN before evaluation and was blinded to the origin sources. Each video was initially assessed by three primary raters (JS, QZ, and ZS). If their scores were unanimous, that score was final. For scores within a one-point range, the principle of majority determined the final score (e.g., mDISCERN scores of 1, 2, 2 and GQS scores of 3, 3, 2 would result in mDISCERN=2, DQS=3). If the score discrepancy exceeded one point (e.g., 2, 3, 4), two secondary raters (FS and RX) reassessed the video. If discrepancies remained, a tertiary grader (YZ) then held a final discussion to reach a consensus. Raters recorded their scores in a scoring spreadsheet; primary raters entered scores for all videos, while secondary and tertiary raters only recorded scores for videos they reviewed.

In this study, we adopt the Fleiss Kappa coefficient to measure consistency among three or more scores, whereas Cohen’s Kappa is utilized for the measurement between two scores. To validate the reliability of our analysis results and conclusions, we calculated the Fleiss kappa coefficient and its p-value to gauge the consistency of the scores given by the three primary raters. To further explore the difference in video ratings between professional physicians and the general audience, we engaged three raters (QL, SY, and ZX) with higher education but not medical expertise to rate the videos based on the same criteria. Then, we computed the Cohen’s Kappa between the final scores from professional raters and those of the non-professional raters.

## Results

### Basic Characteristics of Videos

We used the keyword “dietary weight loss” to retrieve videos on three platforms respectively. During the retrieval process, we found that there were only about 90 videos related to “dietary weight loss” in TikTok search, but there were 150 video recommendations using “ dietary weight loss scheme” as the keyword search. Finally, after the screening, the top 100 videos will be retained in BiliBili, TikTok, and Kwai. The basic information of the video is presented in Table 3. Note that the video duration on BiliBili was significantly longer than that on TikTok and Kwai (p<0.01). In contrast, TikTok and Kwai had higher numbers of likes, collections, and comments, likely due to their shorter video formats (p<0.01).

**Table 3:** Characteristics of videos collected from three platforms.

| Variable | BiliBili(med) | TikTok(med) | Kwai(med) |
| --- | --- | --- | --- |
| GQS | 2 ( 0-4 ) | 2 ( 1-3 ) | 2 ( 1-3 ) |
| mDISCERN | 2 ( 0-5 ) | 2 ( 0-3 ) | 2 ( 0-3 ) |
| Duration(s) | 325(12-13200) | 72(6-384) | 70(6-474) |
| Likes(k) | 1.845(0.04-379) | 60.5(0.04-1765) | 108.5(2-1253) |
| Save(k) | 1.153(0.002-5.9) | 9.5(0.004-70) | 34(1.3-441) |
| Comment(k) | 0.228(0.009-4.421) | 2.2(0.018-208) | 4.6(0.02-84) |

Figure 2 illustrates the distribution of the video source and content type on three platforms. Considering the theme of our video collection is dietary weight loss, the content of the video mostly starts from the creator themselves, sharing their weight loss experience or knowledge, with the remainder being advice from doctors on how to lose weight healthily. Accordingly, we categorized the collected videos into two groups: those published by professional doctors and those not. Among all videos included in this study, BiliBili had 18 videos from professional doctors, TikTok had 5, and Kwai had 20.

**Figure 2:**
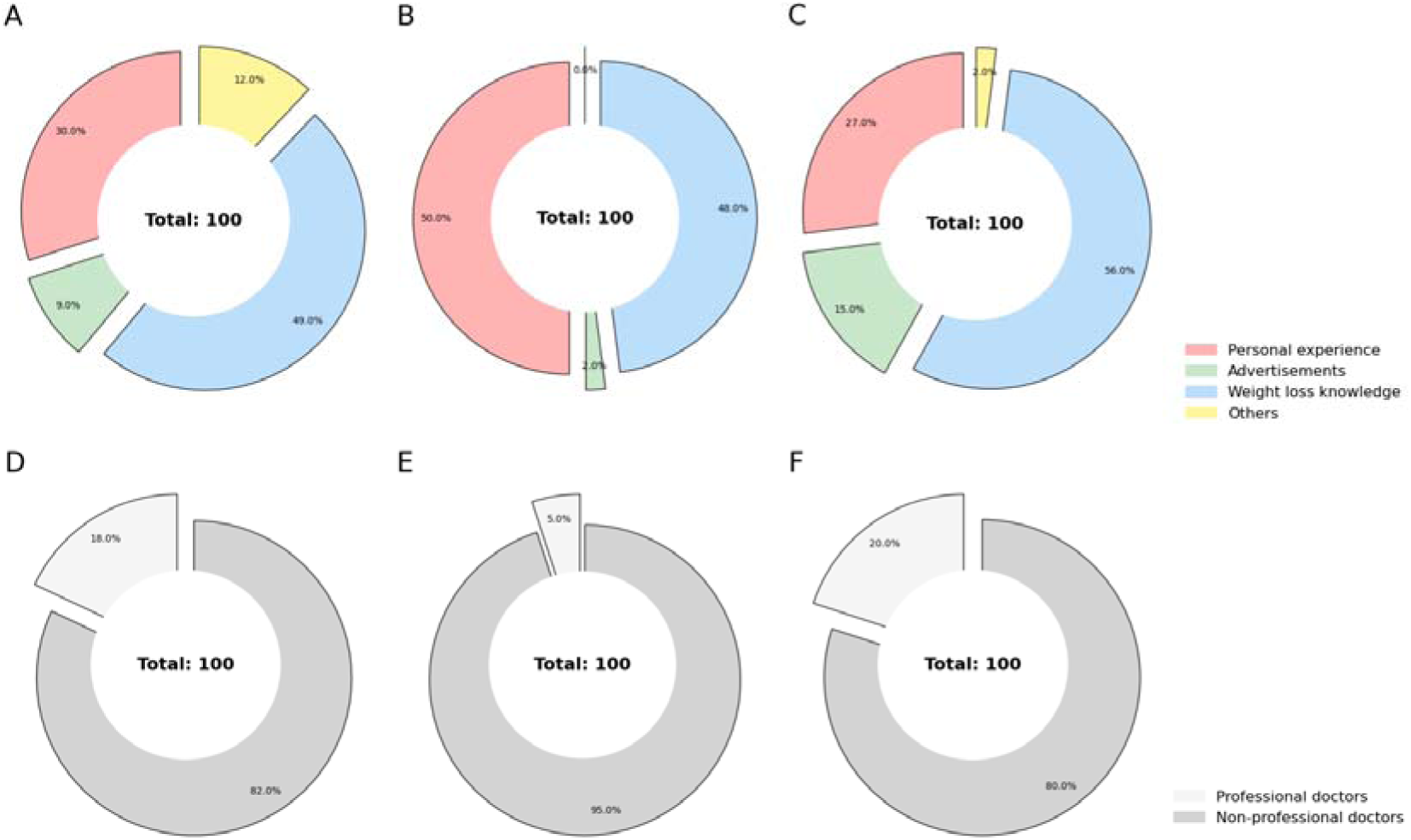
Percentage of videos on dietary weight loss with different contents and from different sources on different platforms. The first row presents the different contents of video on (A) BiliBili, (B) TikTok, and (C) Kwai, respectively. The second row presents the different sources of video on (D) BiliBili, (E) TikTok, and (F) Kwai, respectively.

Additionally, videos have been classified into four distinct content categories: personal experiences, weight loss knowledge, advertisements, and others. The number of videos focusing on personal experiences on BiliBili, TikTok, and Kwai were 30, 50, and 27, respectively. The number of videos dedicated to weight loss knowledge totaled 49, 48, and 56 for each platform. The count of videos that contain advertisements were 9, 2, and 15, while the remaining videos totaled 12, 0, and 2 for BiliBili, TikTok, and Kwai, respectively.

### Video Quality and Reliability Assessments

#### Overall Comparison across Platfroms

As shown in Figure 3, after three rounds of rating, the average GQS scores for dietary weight loss videos on BiliBili, TikTok, and Kwai are 2.04, 1.81, and 1.7, respectively, while the average mDISCERN scores are 2.01, 1.81, and 1.73. The median scores for both GQS and mDISCERN across the three platforms are 2. Although BiliBili’s GQS and mDISCERN scores are higher than those of Kwai and TikTok (GQS: p<0.01, p=0.02; mDISCERN: p=0.02, p=0.08), none of the platforms have exceeded a score of 3, indicating that the quality and reliability of dietary weight loss videos on BiliBili, TikTok, and Kwai are all quite poor. This result may be attributed to insufficient oversight of the scientific rationality and authenticity of their video content on these platforms.

**Figure 3.**
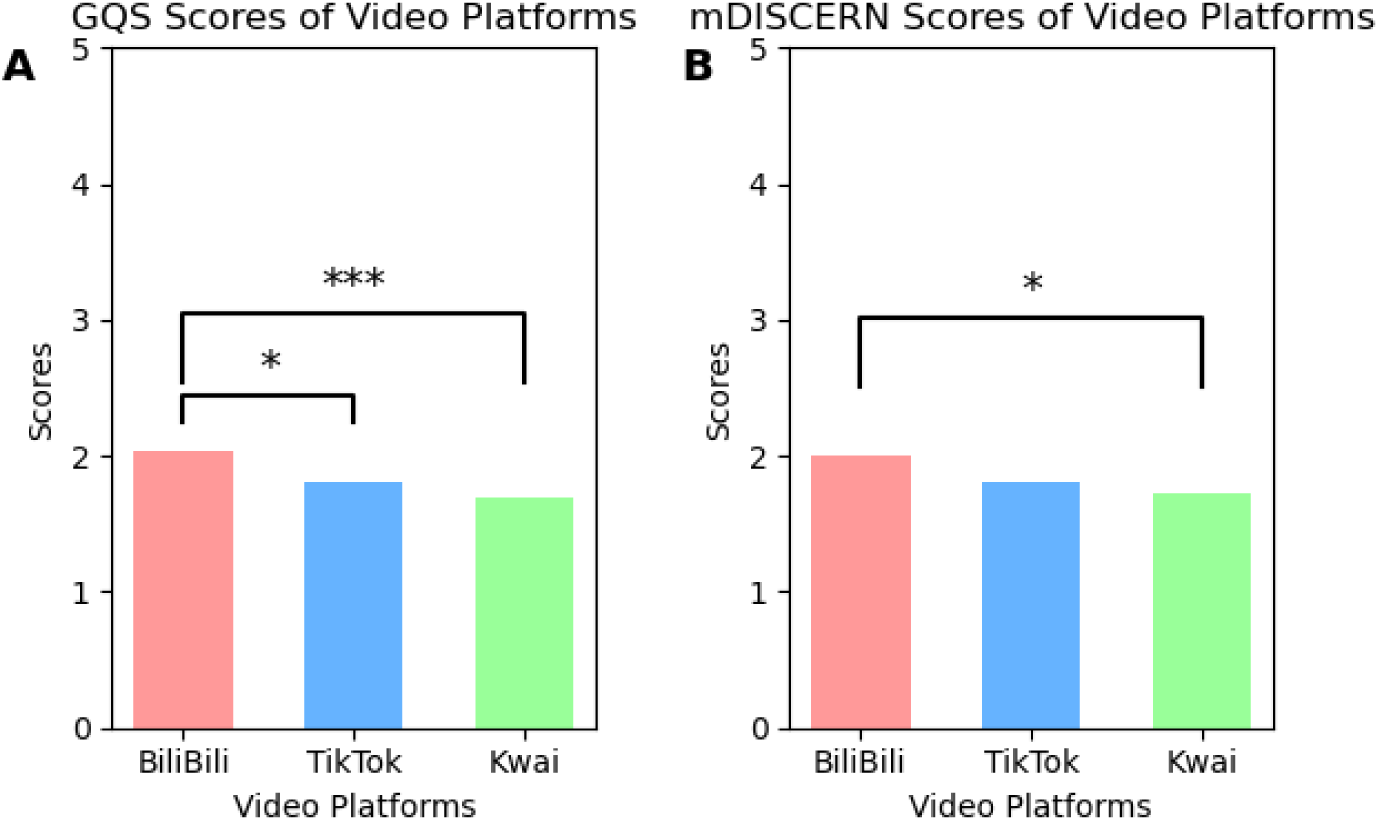
Comparison of (A) GQS and (B) mDISCERN scores determined by professional raters from three video platforms. *p<.05, **p<.01, ***p<.001.

In the first round of rating for dietary weight loss videos on BiliBili, the average GQS scores assigned by the three doctors were 2.08, 2.26, and 2.32, with medians of 2, 2, and 2, respectively, while the average mDISCERN scores were 1.95, 2.18, and 2.24, also with medians of 2, 2, and 2. For TikTok videos, the average GQS scores were 1.90, 1.84, and 2.02, with medians of 2, 2, and 2, and the average mDISCERN scores were 1.81, 1.84, and 2.48, with medians of 2, 2, and 3. On Kwai, the average GQS scores were 1.87, 1.63, and 1.63, all with medians of 2, while the average mDISCERN scores were 1.86, 1.73, and 2.17, with medians of 2, 2, and 2. Table 4 provides a detailed comparison of GQS and mDISCERN scores among the three primary raters, along with the corresponding Fleiss Kappa coefficients. Notably, all Fleiss Kappa coefficients exceeded 0.90, indicating strong inter-rater reliability during the first round of scoring.

**Table 4.** Doctor’s first round scoring results.

|  | GQS |  |  | mDISCERN |  |  |
| --- | --- | --- | --- | --- | --- | --- |
|  | BiliBili | TikTok | Kwai | BiliBili | TikTok | Kwai |
| Rater #1 | 2.08 | 1.90 | 1.87 | 1.95 | 1.81 | 1.86 |
| Rater #2 | 2.26 | 1.84 | 1.63 | 2.18 | 1.84 | 1.73 |
| Rater #3 | 2.32 | 2.02 | 1.63 | 2.24 | 2.48 | 2.17 |
| Fleiss Kappa | 0.90 | 0.95 | 0.98 | 0.90 | 0.93 | 0.94 |

Table 5 compares the average GQS and mDISCERN scores assigned by professional and non-professional raters across the three platforms. The results indicated poor agreement between experts and the general audience regarding the quality and reliability of dietary weight loss videos, with the highest Cohen’s Kappa coefficient being less than 0.2.

**Table 5.** Comparison of scores between professional and non-professional raters.

|  | GQS |  |  | mDISCERN |  |  |
| --- | --- | --- | --- | --- | --- | --- |
|  | BiliBili | TikTok | Kwai | BiliBili | TikTok | Kwai |
| Professional | 2.04 | 1.81 | 1.70 | 2.01 | 1.81 | 1.73 |
| Non-professional | 2.10 | 2.29 | 2.77 | 1.67 | 1.91 | 2.41 |
| Cohen's Kappa | 0.19 | 0.05 | 0.04 | 0.13 | 0.12 | 0.03 |

#### Comparison Under Different Categories

On BiliBili, the dietary weight loss videos published by professional doctors scored lower than those published by non-doctors in both GQS and mDISCERN scores (all P<0.05). On TikTok, the dietary weight loss videos published by professional doctors scored better than those published by non-doctors in both GQS and mDISCERN scores (GQS: P=0.43; mDISCERN: P<0.05). On Kwai, the videos published by professional doctors did not perform as well as those by non-doctors in GQS scores (P=0.68), but in mDISCERN scores, the videos published by professional doctors outperformed those by non-doctors (P=0.02). Detailed comparison is shown in Table 6.

**Table 6.** Comparison of scores for videos from doctor and non-doctor creators.

|  | GQS |  |  | mDISCERN |  |  |
| --- | --- | --- | --- | --- | --- | --- |
|  | BiliBili | TikTok | Kwai | BiliBili | TikTok | Kwai |
| Doctor | 1.56 | 2.00 | 1.65 | 1.50 | 2.60 | 2.05 |
| Non-doctor | 2.13 | 1.80 | 1.71 | 2.10 | 1.77 | 1.65 |

As shown in Table 7, videos focused on weight loss knowledge received the highest GQS and mDISCERN scores across all platforms. Personal experience videos ranked second, except for the mDISCERN scores on Kwai, where advertisement videos held the second-highest position. As TikTok did not have videos int the other category, the corresponding items in the table are marked by “/”. Figure 4 visualizes the comparison among four content categories with significance. Although weight loss knowledge videos were highest in all groups, it had no significant difference from the second place.

**Figure 4:**
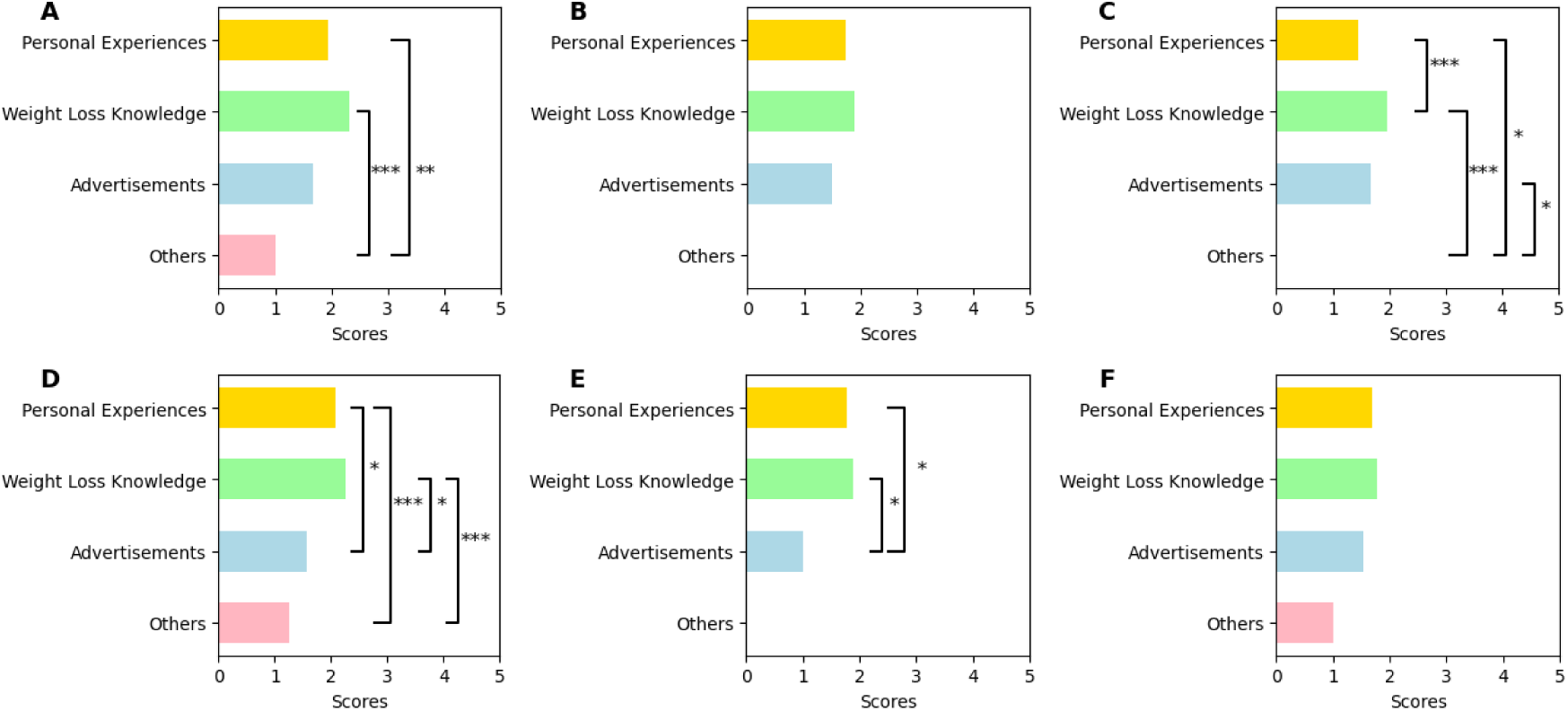
Comparison of video scores in different content categories. The first row presents the mDISCERN scores of (A) BiliBili, (B) TikTok, and (C) Kwai, respectively. The second row presents the GQS scores of (D) BiliBili, (E) TikTok, and (F) Kwai, respectively. *p<.05, **p<.01, ***p<.001.

**Table 7:** Comparison of score for videos with different contents. The highest score of each column is in italics. “/” means that no samples in the corresponding category.

|  | GQS |  |  | mDISCERN |  |  |
| --- | --- | --- | --- | --- | --- | --- |
|  | BiliBili | TikTok | Kwai | BiliBili | TikTok | Kwai |
| Personal experiences | 2.07 | 1.78 | 1.70 | 1.93 | 1.74 | 1.44 |
| Weight loss knowledge | *2.26* | *1.88* | *1.77* | *2.31* | *1.90* | *1.95* |
| Advertisements | 1.56 | 1.00 | 1.53 | 1.67 | 1.50 | 1.67 |
| Others | 1.25 | / | 1.00 | 1.00 | / | 0.00 |

#### Correlation Analysis

Since the collected data are not normally distributed, we utilized the Spearman correlation coefficient to analyze the relationships between different variables. Positive correlations were found between the GQS score and video duration (r=0.41, P<0.01), the mDISCERN score and video duration (r=0.32, P<0.01), the number of likes and the number of favorites (r=0.90, P<0.01), the number of likes and the number of comments (r=0.92, P<0.01), as well as the number of favorites and the number of comments (r=0.86, P<0.01). Negative correlations were observed between the GQS score and the number of likes, favorites, and comments (r=-0.18, P<0.01; r=-0.10, P=0.085; and r=-0.17, P<0.01), the mDISCERN score and the number of likes, favorites, and comments (r=-0.12, P=0.04; r=-0.02, P=0.77; and r=-0.13, P=0.03), and video duration with the number of likes, favorites, and comments (r=-0.36, r=-0.22, r=-0.32, all P<0.01).

## Discussion

### Principal Findings

In this cross-sectional study, we collected and reviewed videos on the topic of dietary weight loss from the three most popular video platforms in China, namely BiliBili, TikTok, and Kwai. Based on data such as the number of likes and favorites, videos on TikTok and Kwai are more popular, while videos on BiliBili tend to have a longer duration compared to the other two platforms. We also assessed the quality and reliability of the videos using the mDISCERN and GQS scales. The mDISCERN and GQS scoring results indicated that BiliBili’s scores for both GQS and mDISCERN are higher than those of Kwai and TikTok (GQS: P=0.02, P=0.001; mDISCERN: P=0.11, P=0.03). Overall, the quality and reliability of videos of dietary weight loss across these three platforms were unsatisfactory, likely due to the inadequate oversight of the scientific accuracy and authenticity.

### Video source, Video Content and Video Quality

First, we observed a clear contrast in the rankings of scores between professional and non-professional raters across the three platforms. For professional raters, the GQS and mDISCERN scores were ranked from highest to lowest as BiliBili, TikTok, and Kwai. In contrast, non-professional raters ranked the platforms from highest to lowest as Kwai, TikTok, and BiliBili. This observation implied that general audience and experts might pay attention to different aspects when they watched health related videos. Video creators should notice this distinguishment and balance the attractiveness and correctness of their content.

Second, we found that videos from TikTok published by professional doctors scored higher on both the GQS and mDISCERN scales compared to those by non-doctors.

However, on the other two platforms, videos by professional doctors did not outscore those by non-doctors, which is contrary to previous research findings [19]. A possible explanation is that videos published by the non-doctor community tend to be more personal, sharing their own experiences and insights on dietary weight loss, while videos by professional doctors were more advisory in nature, presenting knowledge that was more generalized and less “personalized.” Moreover, the theme selected for this experiment, dietary weight loss, did not require a strong professional medical background compared to popular science videos on cancer or liver disease; every video creator had their own experiences and insights on weight loss. Therefore, professional doctors should further optimize their video content, tailoring different dietary weight loss videos for different audiences, thereby conveying more beneficial health knowledge to society.

Third, we found that if the video content is related to weight loss knowledge, it scores higher on the GQS and mDISCERN scales than the other three types of content. Regardless of the content, BiliBili videos scored higher than those on Kwai and TikTok. Personal experiences with dietary weight loss might spread misinformation, such as losing weight by eating only a cucumber at night or not eating at all and how much weight was lost as a result. In contrast, videos sharing weight loss knowledge were based on scientific evidence, such as the “16+8 diet” or “ketogenic diet,” naturally scoring higher than other types of content.

### The Correlation Between Video Quality and Video Characteristics

We found that the quality of videos is positively correlated with video duration, which was consistent with previous research findings [31]. As the length of the video increases, creators could convey more information to the audience, which to some extent enhances the reliability and quality of the video. We also discovered that the quality and duration of the video was negatively correlated with likes, favorites, and comments, aligning with many previous studies [21, 22, 32]. This suggested that the audience had poor capability to distinguish between high-quality and low-quality videos.

As short video users usually watch content during their leisure time, shorter videos typically have higher completion rates and reach a larger audience. Additionally, the push mechanisms on short video platforms mean that videos with more likes are likely to rank higher and gain more exposure. Consequently, shorter, lower-quality videos tend to be more popular, which exacerbates the gap between video quality and popularity.

Therefore, video creators need to strike a balance between video duration, content richness, and reliability. Namely, how to convey high-quality health and medical science videos to the audience in a short time using concise and easily understandable language.

### Evaluation of Quantitative Scoring Tools

We utilized the mDISCERN and GQS to assess the reliability and quality of information in the videos. The mDISCERN is a 5-point scale adapted from DISCERN, which was originally developed to enable patients to judge the quality of written information [25, 33]. However, it has been noted that DISCERN is not suitable for assessing the reliability of videos [28]. The GQS, proposed by Bernard et al., was initially designed to assess the quality of information on websites but has since been widely adopted for evaluating video content [26, 30], with its effectiveness for video quality assessment supported by previous studies [30, 34, 35]. Nevertheless, both the GQS and mDISCERN have certain limitations. They do not evaluate the content of the video comments, focusing more on the textual content of the videos, which we believe is a direction for improving future video assessment tools.

### Practical significance

Over the past forty years, more than half of Chinese adult population has become overweight or even obese [36]. Obesity can lead to a range of metabolic disorders such as hypertension, hyperglycemia, and hyperuricemia, which pose a serious threat to public health. A multitude of evidence suggests that a weight loss of 5-15% can significantly improve metabolic disorders and reduce the risk of cardiovascular diseases and other weight-related comorbidities [37, 38]. With the continuous development of science and technology, the primary source from which people obtain health information has gradually shifted from medical books to the internet. However, due to the lack of effective regulatory mechanisms, the quality of information on the internet is uneven, and some videos mislead consumers by providing inaccurate information [39, 40]. Therefore, it is essential to employ scientific scoring tools to assess diet and weight loss videos. High-quality health videos can raise awareness of obesity and contribute positively to public health education. In response to this need, the Chinese government has issued the world’s first health promotion guide [41]. Video platforms should take proactive steps by establishing dedicated medical sections, forming expert review teams to assess medical content, and refining video ranking and recommendation algorithms to ensure high-quality medical videos are more accessible. Additionally, medical professionals must be accountable for the accuracy and reliability of the videos they produce.

### Advantages and Limitations

To the best of our knowledge, our research is the first to assess the reliability and quality of internet videos on the topic of dietary weight loss from Chinese videos platforms. We avoided the limitation of studying only a single video platform by selecting the most popular short video platforms used by Chinese users. BiliBili, primarily used by younger audiences, along with TikTok and Kwai, which cover audiences of all ages and cultural backgrounds, make our study more reliable and comprehensive. We employed the GQS and the mDISCERN tool to assess videos, providing a thorough and systematic evaluation of the quality and reliability of the videos. In addition, the doctors who participated in the assessment have extensive clinical experience in the field of diet and weight loss. To minimize subjective bias in video scoring, we adopted a three-tier scoring system in the assessment process.

Videos with score discrepancies at the first level were reviewed by the second-level raters, and those with remaining discrepancies were determined based on a discussion held by the third-level rater.

Previous studies have shown that the top 100 videos can represent the overall quality of the field’s videos [22, 42], but as time goes on, the top 100 videos on video platforms will also change. Our study focused videos from Chinese platforms and did not assess or compare contents from non-Chinese videos, which limits the generalizability of our findings to international video platforms.

## Conclusions

We collected 100 videos on the theme of dietary weight loss from BiliBili, TikTok and Kwai, and evaluated their reliability and quality using the GQS and mDISCERN tools. After three rounds of scoring, we found that while the GQS and mDISCERN scores for BiliBili were higher than those of TikTok and Kwai, the final scores of the three platforms did not exceed 3 points, which indicated that the video quality of the three video platforms on the theme of dietary weight loss was low. Video social media platforms should establish relevant policies to supervise and review the publication of medical science popularization videos, in order to prevent users from receiving incorrect medical knowledge. At the same time, medical practitioners are encouraged to actively create high-quality medical science popularization videos, providing reliable sources of information for public health education.

## Declarations

## Ethics approval and consent to participate

Not applicable

## Consent for publication

Not applicable

## Availability of data and materials

The datasets used or analyzed during the current study are available from the corresponding author on reasonable request. Please contact the

## Conflicts of Interest

The authors declare that they have no competing interests

## Funding

Not applicable

## Authors’ contributions

YC and YX conceived the study. XC and HY developed research design. QL, SY, and ZX were responsible for collecting and filtering the online videos. YZ, QZ, FS, RX, ZS, and JS were responsible for reviewing and scoring the videos. SZ and LY analyzed the data. XC and SZ wrote the original draft. YC, HY, and YX reviewed and edited the manuscript. All the authors approved the final draft for submission.

## Data Availability

All data produced in the present study are available upon reasonable request to the authors

## Acknowledgements

Not applicable

## Authors’ information

SZ and XC are co-first authors.

## Notes

### Competing Interest Statement

The authors have declared no competing interest.

### Funding Statement

This study did not receive any funding

### Author Declarations

All data were sourced from publicly available videos on BiliBili, TikTok, and Kwai, ensuring no personal privacy issues were involved. As the study did not engage directly with users, it did not require ethical review.

